# Comparing the predictive power of machine learning and semi-mechanistic models of endemic measles dynamics

**DOI:** 10.1101/2022.05.27.22275695

**Authors:** Max SY Lau, Alex Becker, Lance Waller, Jessica Metcalf, Bryan Grenfell

## Abstract

Measles is one the best-documented and most-mechanistically-studied non-linear infectious disease dynamical systems. However, systematic investigation into the comparative performance of traditional mechanistic models and machine learning approaches in forecasting the transmission dynamics of this pathogen are still rare. Here, we compare one of the most widely used semi-mechanistic models for measles (TSIR) with a commonly used machine learning approach (LASSO), comparing performance and limits in predicting short to long term outbreak trajectories and seasonality for both regular and less regular measles outbreaks in England and Wales (E&W) and the United States. First, our results indicate that the proposed LASSO model can efficiently use data from multiple major cities and achieve similar short-to-medium term forecasting performance to semi-mechanistic models for E&W epidemics. Second, interestingly, the LASSO model also captures annual to biennial bifurcation of measles epidemics in E&W caused by susceptible response to the late 1940s baby boom. LASSO may also outperform TSIR for predicting less-regular dynamics such as those observed in major cities in US between 1932-45. Although both approaches capture short-term forecasts, accuracy suffers for both methods as we attempt longer-term predictions in highly irregular, post-vaccination outbreaks in E&W. Finally, we illustrate that the LASSO model can both qualitatively and quantitatively reconstruct mechanistic assumptions, notably susceptible dynamics, in the TSIR model. Our results characterize the limits of predictability of infectious disease dynamics for strongly immunizing pathogens with both mechanistic and machine learning models, and identify connections between these two approaches.

## Introduction

Mechanistic and semi-mechanistic models have been foundational in developing an understanding of the spread of infectious diseases in human and wildlife populations. These models approximately depict how pathogen transmission is shaped by population dynamics (e.g., how transmission is reduced by herd immunity). Such modeling approaches are essential for understanding the natural history of pathogens transmission and providing insights into designing effective control strategies. While models such as the Susceptible-Infected-Recovered framework are mechanistically well-understood, calibrating them against stochastic, and often partially unobserved, incidence or mortality data is a steep statistical challenge. The primary focus of mechanistic models has been *understanding and characterizing* the natural history of transmission. In contrast to their mechanistic counterparts, implementations of statistical and machine learning techniques in infectious disease modeling have primarily focused on improving *forecasting* accuracy without the explicit aim of inferring transmission dynamics. Such approaches have grown in popularity in recent years^1–5^, and they also have a long pedigree in terms of using statistical approaches to study measles dynamics ^6,7^.

Patterns of pre- and post-vaccination measles incidence are among the most well-documented, and well-studied, non-linear systems in ecology. A suite of analyses using deterministic and stochastic (semi-) mechanistic models have illuminated how the interplay between seasonal forcing and susceptible recruitment shape dynamics in large urban populations ^8^, ranging from simple limit cycles to coexisting attractors ^8,9^, and even chaos with the domination of stochastic extinction in small highly vaccinated populations^10,11^. A focus of previous analysis has been detailed weekly spatio-temporal notifications of measles from England and Wales (E&W), interpreted with the TSIR model and other inferential approaches, notably particle filtering ^12,13^. While partially mechanistic approaches for measles dynamics are being explored ^14–16^, a more comprehensive comparison between mechanistic and fully statistical approaches is still lacking. Such comparisons would yield insight into the choice of most appropriate modeling techniques given different patterns of data. Measles is an excellent test bed for these questions, given that we have both rich historical notification time series and successful applications of mechanistic and semi-mechanistic models.

In this paper, we explore and compare forecasting capability of these two contrasting approaches for both regularly periodic and relatively irregular recurrent measles epidemics in England and Wales between 1944-1994 and in the US between 1932-45. We consider both a stochastic semi-mechanistic TSIR model and a fully-statistical model using a popular machine learning (ML) approach (Least Absolute Shrinkage and Selection Operator, the LASSO).

Our results suggest that the proposed LASSO model, compared to the TSIR model, can efficiently use data from multiple major cities and achieve similar short-to-medium term forecasting performance for more regular measles outbreaks in E&W during the pre-vaccination era (1944-1964). Strikingly, even when trained solely on data with an annual cycle, forecasts in our LASSO framework capture the characteristic annual to biennial bifurcation 1950 driven by a decline in birth rates. When important demographic information (such as the birth rate data) is not included, the LASSO model still performs reasonably well, likely due to the fact these dynamics may have been implicitly incorporated within the approach (see Models and Methods). LASSO may also outperform the TSIR for predicting less-regular dynamics such as those observed in major cities in the US between 1932-45. Although both approaches capture short-term forecasts, accuracy suffers for both methods as we attempt longer-term predictions in highly irregular, post-vaccination epidemics in E&W. Overall, our results show that fitting a LASSO model may both qualitatively and quantitatively rediscover major mechanistic assumptions in the TSIR model. These insights inform the limits of predictability, and the connections of both approaches in infectious disease dynamics for fully-immunizing pathogens.

## Results

### Forecasting measles outbreaks in England and Wales

Measles dynamics pre- and post-the introduction of mass-vaccination program are particularly well-documented from historical notifications in England and Wales ^17^. Before widespread vaccination in the late 60s, measles epidemics in E&W were characterized by highly regular periodic (often biennial) cycles in large cities (see Figure 1 showing outbreaks in London). Analyses often focus on populations at, or above, the Critical Community Size (i.e., the endemic threshold, CCS) of approximately 300,000 individuals ^18^. We follow this precedent here and focus on modeling eight major cities, each with a population above the CCS: namely, London, Liverpool, Birmingham, Manchester, Nottingham, Bristol, Leeds and Sheffield. Although dynamics were highly-consistent in the pre-vaccine era, the introduction of measles vaccination in 1968 led to reduction in both epidemic size and regularity.

**Figure 1:**
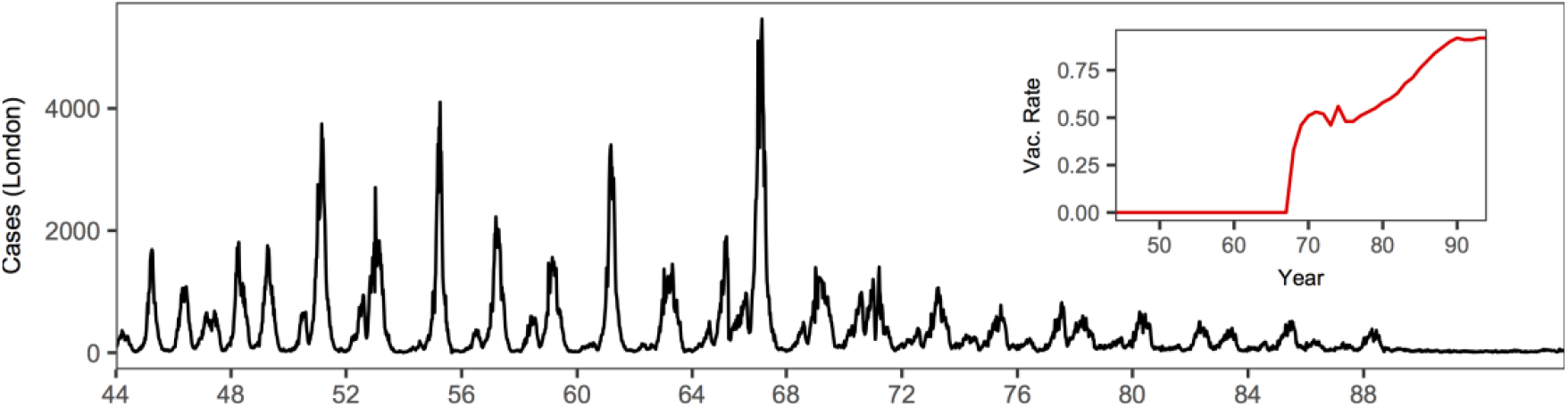
Long-term dynamics of measles in London. Bi-week incidence of measles cases in London (1944-94) and time series of vaccination. Following the widespread vaccination in the late 60s, the epidemics shifted from highly regular cycles to largely irregular dynamics.

We fit both the LASSO model and the TSIR model (see *Models and Methods*) to the measles epidemics in those major cities in the pre-vaccination era, using the data from 1944-51 for model training. We illustrate our findings by comparing two approaches in predicting epidemics in London. Figure 2 and Figure S1 shows that the TSIR model can predict the epidemic reasonably accurately in short to medium-term. In particular, while both approaches are able to capture the trends of the epidemic trajectories, their accuracy generally decline from 8-biweeks in the future. Although direct inference of a single location (e.g., London) is relatively straightforward in the TSIR framework, incorporating incidence from multiple locations is challenging ^9,17^. The LASSO model, when incorporating all the available incidence data from the major cities, is able to achieve similar performance (Figure S3 also shows that TSIR clearly outperforms LASSO when only London data is used for model training).

**Figure 2:**
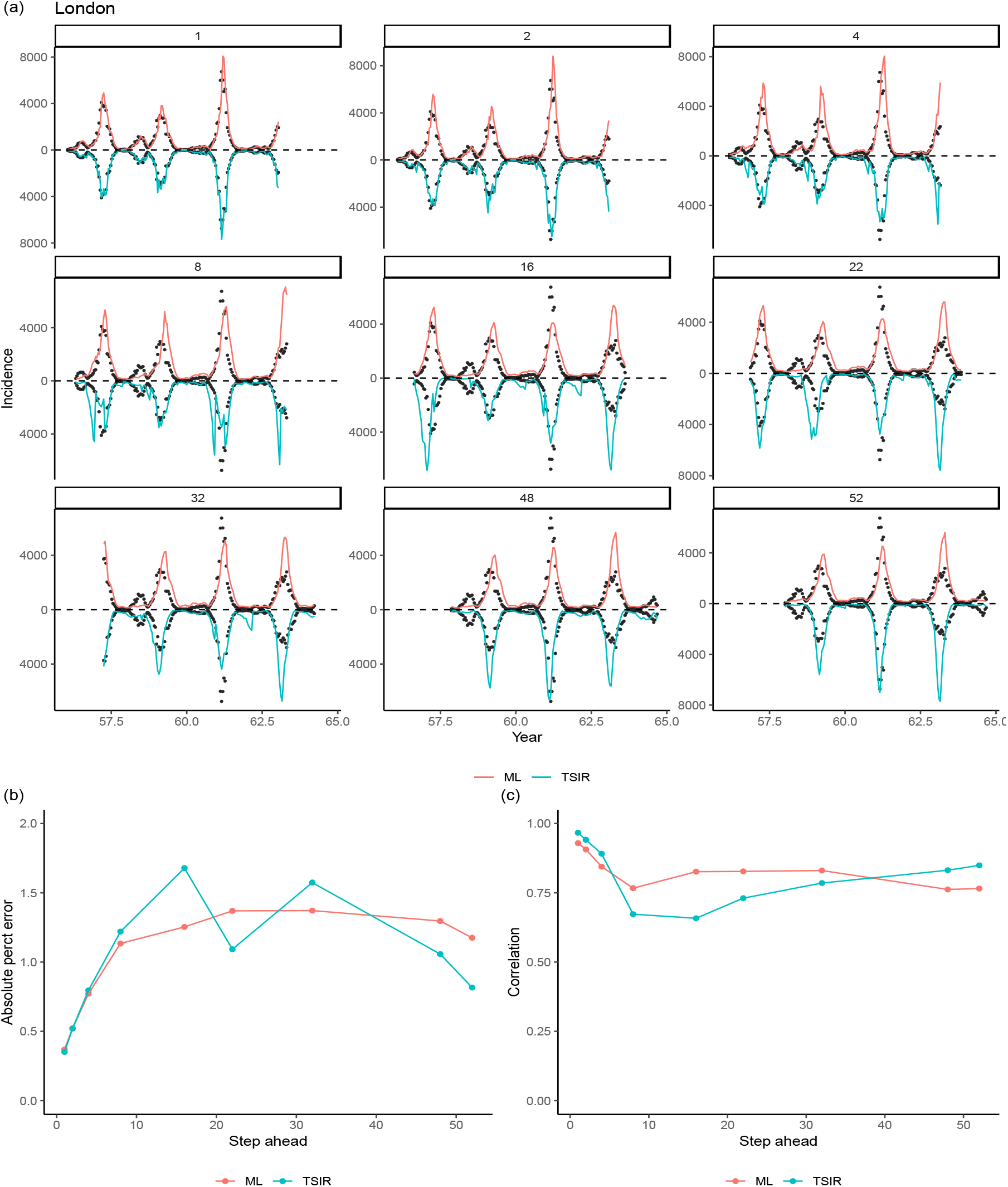
Out of sample predictions for measles in London. A subset of 1 to 52^th^-biweek ahead out-of-sample (i.e., period excluding data in the training set) predictions from our LASSO model and the TSIR model, for pre-vaccination measles epidemics in London from 1944-64. For k^th^- step ahead predictions, the incidence at a particular time point t was predicted using the LASSO model or TSIR model conditional on observations up between time t − k − t_lag_ and t − k where we use t_lag_ = 130 (see Models and Methods). Data between 1944-51 (from 8 places whose average population sizes are greater than the critical community size 300,000) are used to train a model. Dots indicate the observed incidences. (a) Comparisons of predicted epidemic trajectories; (b) Comparisons of the mean absolute prediction error ^19^ in ratio (i.e., the absolute difference between the predicted and observed value divided by the observed value); (c) Comparisons of the trends (summarized by the correlation) between the predicted and observed trajectories. Although predictions made by the TSIR appear to be more volatile, performance of the two approaches are comparable and they both show the general trend of decreasing performance as steps increases.

Birth rate is an important variable for mechanistic models including the TSIR model as it governs the rate of the replenishment of susceptible population (see also Models and Methods). One common feature of the E&W dataset is an observed bifurcation from annual to predominantly biennial dynamics due to the “Baby Boom” in the late 40s. The impact of this demographic shift was particularly strong; dynamics remained biennial until the transient post-vaccination era starting in the late 60s. We found that the LASSO model forecasts identified and captured the bifurcation reasonably well (Figure 3). Interestingly, we find that the ability of LASSO in predicting the bifurcation remains very similar even without the knowledge of births, the primary causal driver of this dynamic perturbation (Figure S2). By heuristically deriving the connections between TSIR and LASSO (see Models and Methods), we show that the impacts of births and susceptibles may have been implicitly incorporated by the LASSO.

**Figure 3:**
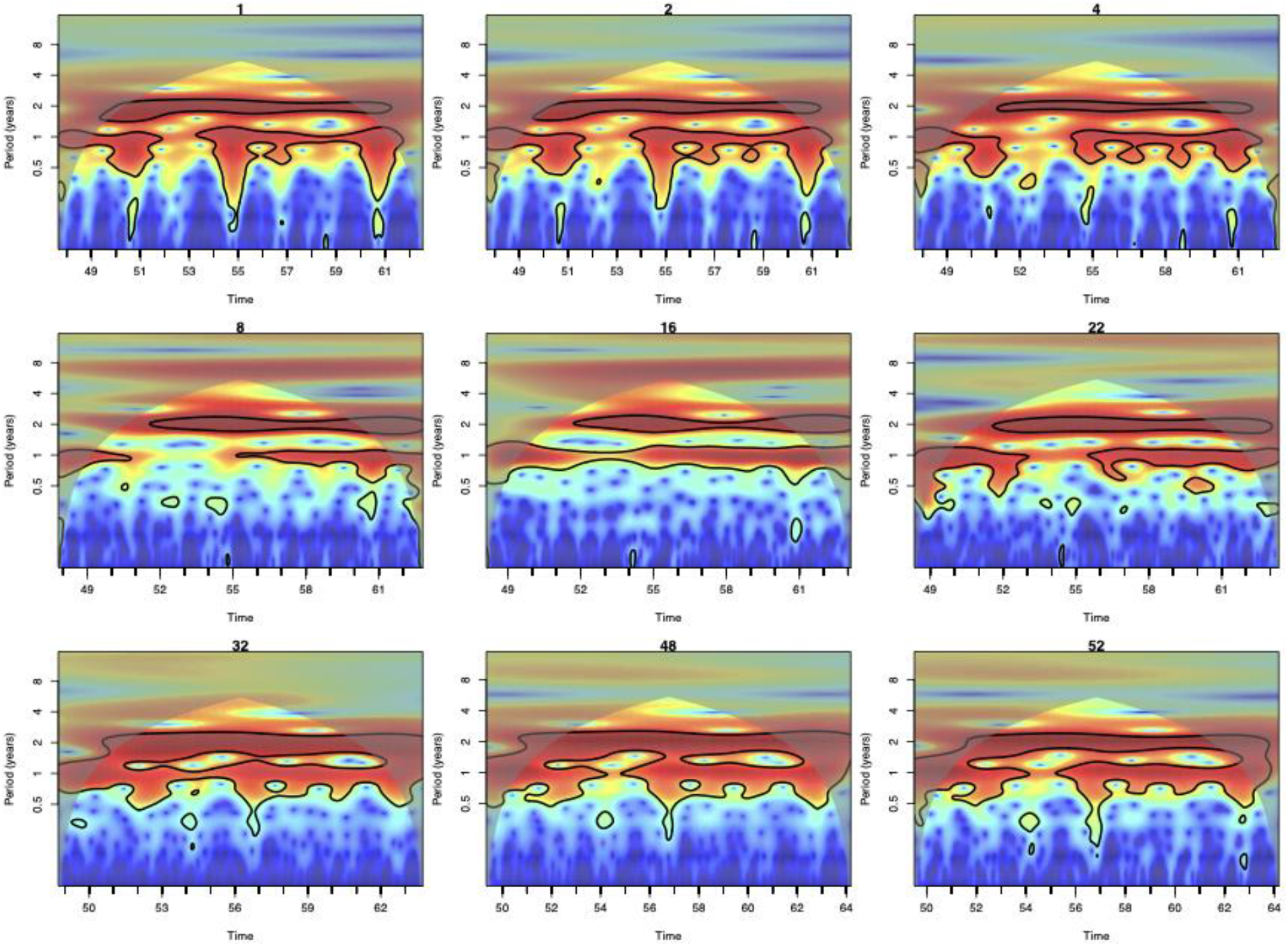
Power spectra of the out of sample predictions for measles in London using the LASSO model. Here we use data between 1944-46 data to train the LASSO models. LASSO successfully captures both measles periodicity and bifurcation timing (1950). For each figure, red indicates regions of identified periodicity. Black contours indicate 95% confidence intervals. Notably, the LASSO model captures the 1950 bifurcation from annual to biennial dynamics starting in 1950.

Turning to the post-1968 vaccine era, both approaches appear to be able to predict highly irregular epidemic trajectories in the short-term among the highly vaccinated populations, but struggle beyond 4-biweek ahead predictions (Figure 4).

**Figure 4:**
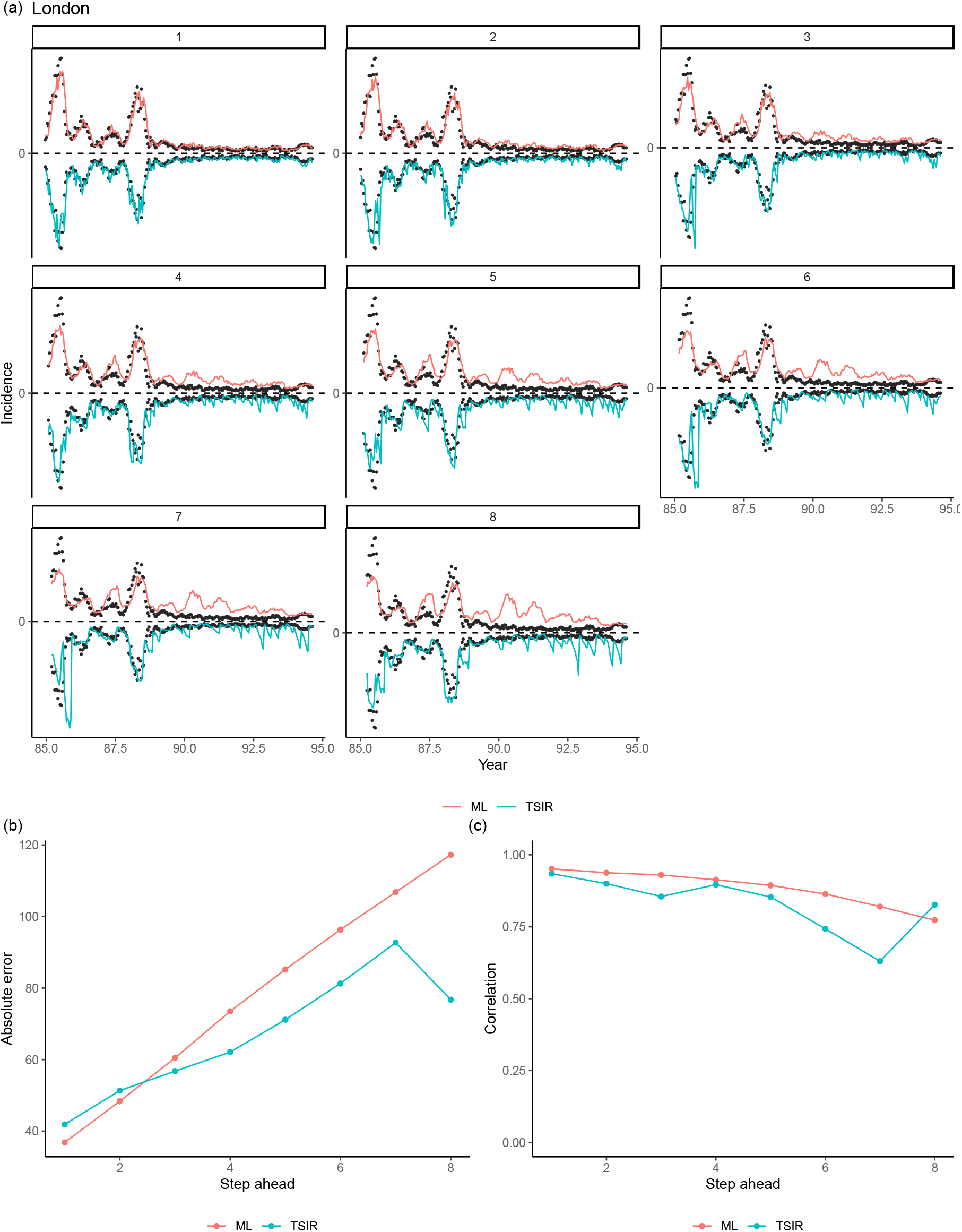
1 to 8^th^-biweek ahead out-of-sample predictions from our LASSO model and the TSIR model, for measles epidemics in London during the vaccination era from 1985-94. Data between 1970-80 (from 8 places whose average population sizes are greater than the critical community size 300,000) are used to train a LASSO model. Note that here we do not compare the prediction error via a ratio (see Figure 2) because observed incidences (the denominator) are often zero in the time period.

### Forecasting pre-vaccination measles outbreaks in the US

In the previous section we illustrated how both the TSIR and the LASSO model can successfully capture key traits (e.g., epidemic size and periodicity) of the observed E&W time series data. However, a rich analytical literature has illustrated the relative dynamic stability of the E&W data (i.e., Lyapunov Exponent < 0) ^8,9^. We now turn our attention to a set of incidence data corresponding to more challenging chaotic dynamics (i.e., Lyapunov Exponent > 0). One such source of data comes from the United States. In contrast to E&W, the US city pre-vaccination measles data exhibit signatures of deterministic chaos^11^. Here, we examine the comparative ability for our LASSO model to predict chaotic dynamics. Fitting both models to the US data, we found the LASSO model may outperform the TSIR model’s ability to capture both the amplitude and trend for the outbreak in New York (Figure 5). Similar results can be found for other US major cities (Figure S4).

**Figure 5:**
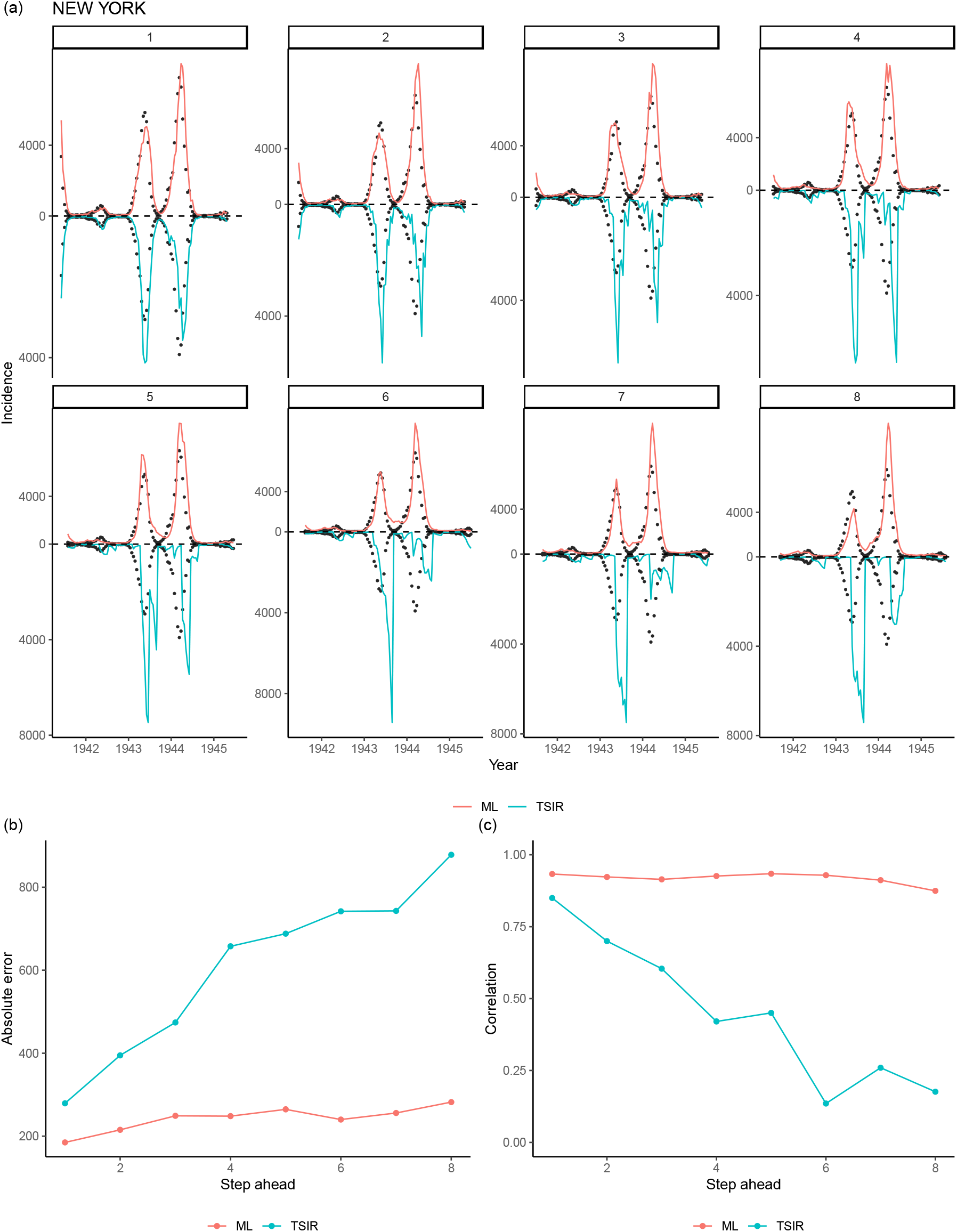
Short-term predictability for irregular dynamics. 1 to 8^th^-biweek ahead out-of-sample predictions from our LASSO model and the TSIR model, for measles epidemics in New York from 1932-45. Data between 1932-40 (from 7 major cities which also have non-missing incidence data during this period) are used to train the LASSO.

### Reconstructing the TSIR mechanism via the LASSO model

Our E&W results show that our LASSO model may reconstruct/rediscover some of the underlying assumptions in the TSIR model. The TSIR model (see also Models and Methods) assumes that the mean incidence at time *t* is governed by the product of incidence and *S*_*t*−1_ the number of susceptibles at the previous time step, i.e.,

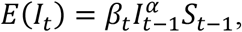

where *β*_*t*_ is a seasonally repeating contact rate with 26 biweekly points per year. The exponent *α* of the TSIR model captures heterogeneities in mixing, which is typically slightly less than 1 (e.g., 0.98). This formulation implies that *I*_*t*_ is expected to be largely positively associated with *I*_*t*−1_, and less so with *I*_*t*−*k*_ for *k* > 1 (as larger *I*_*t*−*k*_ in general lead to larger degree of susceptible depletion and hence smaller *S*_*t*−1_). Secondly, *I*_*t*_ and *I*_*t*−*n*×26_ is also expected to have a positive association due the seasonality assumption embedded in *β*_*t*_. Our LASSO model estimates appear to be able to largely capture these trends (i.e., susceptible depletion and seasonality) implied by the TSIR model (Figure 6). In particular, and our associated LASSO coefficient (of incidence at the 1-biweek lag) is quantitatively capturing the exponent *α* in the TSIR model (Figure 6a). We provide additional insights in the *Models and Methods* regarding how the LASSO model and the TSIR model are interconnected.

**Figure 6:**
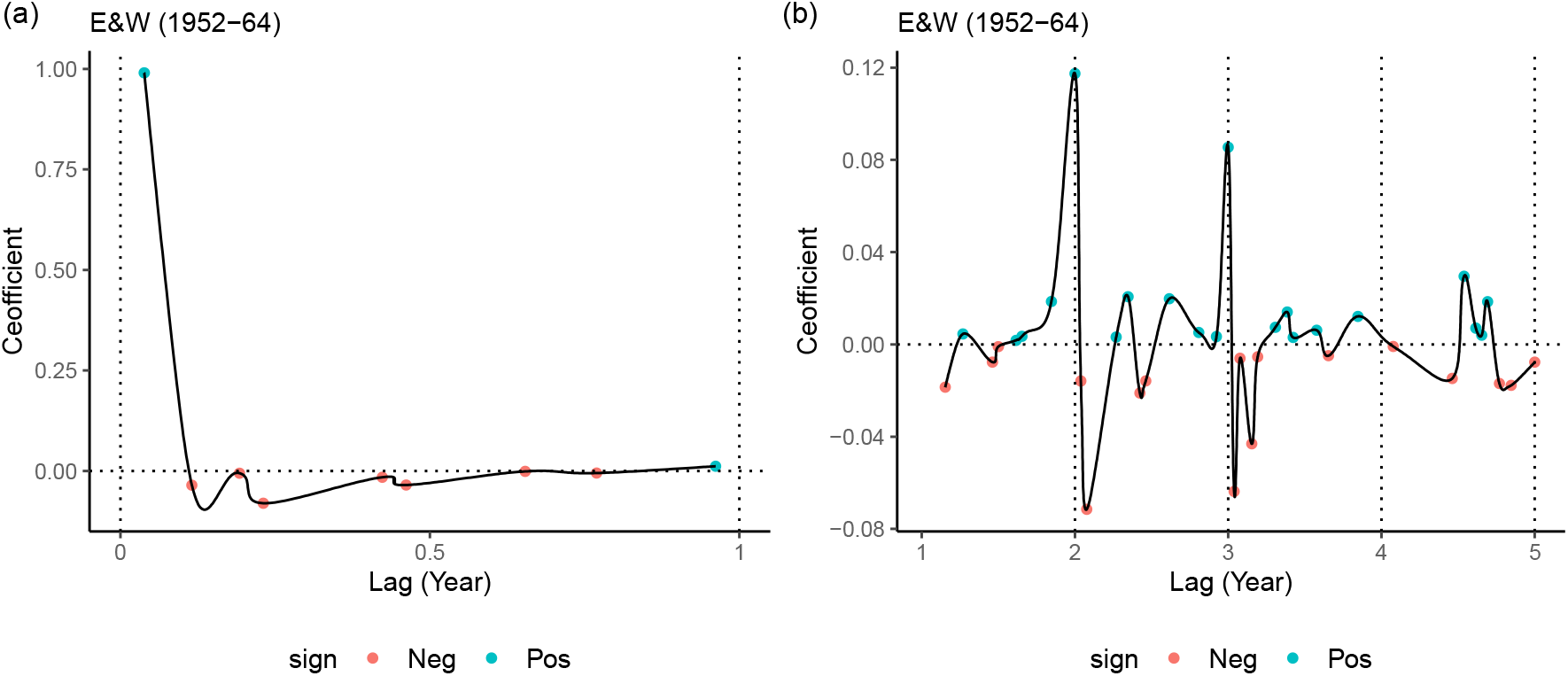
Coefficients associated with lagged incidences in the LASSO model (Equation 4 in Models and Methods), from fitting the one-step ahead model to the pre-vaccination E&W data. Only non-zero LASSO coefficients are shown for clarity. The estimated LASSO model resembles and discovers the TSIR model. (a) Coefficient for the most recent lagged incidence is significantly larger (and positive) and most of the coefficients within one year (26 biweeks) are negative, which is consistent with the TSIR assumptions; moreover, the coefficient of the most recent lag is also quantitatively consistent with the value of the exponent parameter (slightly less than 1) in the TSIR model. (b) Coefficients at 2 and 3-year lags are significantly larger (and positive) compared to coefficients at other lags, consistent with the seasonality assumption of the TSIR model. Coefficient at 1-year does not appear to have a positive association, possibly because it is counterbalanced by the effect of susceptible depletion. Lagged incidences beyond 3 years do not appear to have systematic positive associations.

## Discussion

Transmission of infectious diseases at the population-level is characterized by inherent, and often complex, non-linear dynamics that are driven by intrinsic and extrinsic factors such as infectivity of pathogens, human behaviors and public health interventions, notably variable contact patterns and vaccinations among host populations, and even environmental factors. Mechanistic and semi-mechanistic models provide biologically plausible and directly interpretable frameworks for modeling such complex dynamics. In contrast, machine learning approaches primarily focus on identifying patterns within the data to improve prediction and forecasting; they include no specific mechanistic framing, and often lack biological interpretability. While machine learning approaches have shown success in forecasting complex epidemiological systems (e.g., dengue ^5^), comprehensive and long-term data for these pathogen-human interactions are often lacking, making detailed methodological comparisons challenging. Here, we leverage unique time-series data and a large body of work on semi-mechanistic modeling to develop a full comparison between these approaches using measles as a test case.

Our results indicate that a LASSO-based machine learning model can efficiently leverage the detailed historical measles incidence data from multiple locations in E&W to achieve short to medium-term forecasting accuracy that is comparable to one of the mostly commonly used mechanistic model for measles (the TSIR model). Interestingly, our results show that the LASSO model performs similarly even without the knowledge of births that are required by the TSIR model. This suggests that the correlation/dependence structure between birth and incidence can be “absorbed” by a parsimonious LASSO model that only considers historical incidence to infer changes in temporal patterns without explicit knowledge of the cause of these changes (e.g., here, the impact of births on the underlying susceptible population size). As a result, the LASSO model appears to be able to capture the bifurcation in dynamics in 1950, one of the key properties of the measles outbreaks in E&W, without requiring the data driving the change in pattern. We do, however, find that the LASSO forecasts are comparable to those from TSIR only when all data from the major cities are used for model training (Figure S3). Moreover, while both the LASSO model and the TSIR model do not work well for the highly chaotic dynamics beyond short-term prediction, the LASSO approach may outperform TSIR in the scenarios with a mixture of seasonal and mildly chaotic dynamics (as observed in historical outbreaks in the major cities of US ^11^). Finally, our results also show that the LASSO model can reconstruct/discover the mechanistic assumptions of the TSIR model (Figure 6).

Our results are consistent with recent work which shows that the TSIR model may be discovered by some partially-mechanistic machine learning approaches that consider higher orders of polynomial terms for transmission dynamics ^14^. Compared to their work, our work focuses on out-of-sample prediction (as opposed to focusing on “discovery”). Also, while these approaches require the knowledge of susceptible population (via some pre-processing procedures leveraging TSIR), we do not require reconstructed susceptible population, creating a more explicit test of fully statistical approaches. The effect of susceptible depletion seems to be implicitly captured by the non-positive LASSO coefficient values associated with the more recent lags within the previous year (see Figure 6a and Models and Methods). This feature is likely to explain why the LASSO model may perform reasonably well despite lacking explicit knowledge of births/susceptibles.

This analysis represents an initial step and there are several clear directions for future work: while we compared one of the most successful mechanistic models for measles (the TSIR) with a commonly used machine learning approach (the LASSO), other machine learning approaches (e.g., neural-network based models) may yield different results. In particular, recent theoretical work ^20–22^ has demonstrated excellent predictability for simulated deterministic chaotic systems using a network-based machine learning approach (“reservoir computing”). Such variants could also be extended to and tested on the stochastic measles outbreaks that we consider. Despite this, it is worth noting that the LASSO approach is relatively interpretable compared to many other machine learning approaches, and thus seems a sensible starting point here. Finally, while our preliminary simulation studies (Figure S5) further illustrate the predictive power of the LASSO model for measles epidemics, more extensive simulation studies including the investigations of explicitly leveraging spatial information of the epidemics may be considered for future directions. These investigations may shed light on, for example, how machine learning approaches may best complement mechanistic models for modelling less populated places whose dynamics are known to be more stochastically-driven.

## Models and Methods

### A mechanistic modelling approach: the TSIR model

We model the local measles dynamics using the time-series-Susceptible-Infected-Recovered (TSIR) framework. Balancing births against disease transmission, the TSIR equations are given as

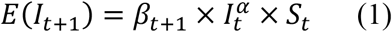

and

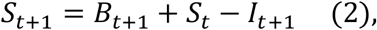

Where *I*_*t*_ and *S*_*t*_ are the number of incident and susceptible individuals in a given biweek *t*, and *B*_*t*_ refers to the number of births in a given biweek, *β*_*t*_ is a seasonally repeating contact rate with 26 values per year, and the exponent *α* (typically slightly less than 1) captures heterogeneities in mixing that were not explicitly modelled by the seasonality ^23,24,14,15^ and the effects of discretization of the underlying continuous time process. The TSIR estimates obtained in this manuscript used the recently developed **tsiR** package ^25^. Specifically, in our analysis, *α* is fixed to be 0.98 ^26^ and a Gaussian process regression is performed between cumulative cases and cumulative births. Parameter estimates were obtained for each location for each time period of interest. A more extensive description of the TSIR fitting process in terms of theory and implementation can be found in ^23,25^.

### A Machine learning approach: the LASSO model

We consider modelling incidence *k*-step ahead of time *t* for place *i* (i.e., *I*_*i,t*+*k*_) as a linear combination of log-transformed historical local incidence and births. Specifically, we consider

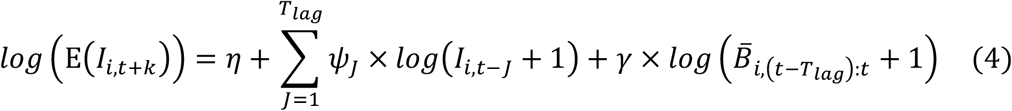

where 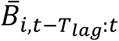 denotes the average of births of the previous *T*_*lag*_ biweeks. We consider bi-weekly data and two-year forecasting windows *k* = 0,1, …, 52 and *T*_*lag*_ = 130. A separate model is fitted for each forecast window *k*, using Least Absolute Shrinkage and Selection Operator (LASSO) regression ^27^. LASSO is a machine learning technique that simultaneously performs estimations of the regression coefficients 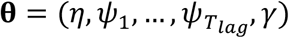 and variable selection by *shrinking* some of the smallest estimated coefficients towards zero. LASSO holds the property of variable selection as it allows a coefficient to be shrunk to *exactly* zero. Compared to the traditional regression technique the Least Squares Estimates (LSE), this shrinkage process has the effect of significantly reducing variance of model prediction and is the key for improving model fit. We also consider a LASSO model without explicit inclusion of the births (the last term in Equation 4).

In particular, LASSO estimates 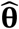 are the values of coefficients that minimize an objective function

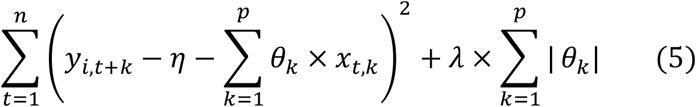

where *y*_*i,t*+*k*_ is the response variable, *p* = 2 × *T*_*lag*_ and *n* is the number of observations. Note that for clarity we have used *θ*_*k*_ to denote the *k*^*th*^ coefficient in θ (not including the intercept *η*) and *x*_*t,k*_ to denote the covariate associated with it. The penalty term 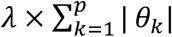 serves as the machinery to allow shrinkage of the coefficient estimates, i.e., the larger the value of *λ*, the greater the effect of shrinkage. Shrinkage significantly reduces variances of predictions, but at the cost of slight increase in bias − and tuning of *λ* is critical for achieving an ‘optimum’ (often measured by the test mean squared error) among this bias-variance trade-off. We used ten-fold cross-validation to identify the optimal value of *λ*.

### Reconstructing the TSIR mechanism via the LASSO model

In this section, we provide additional insights into how our LASSO model can reconstruct/discover a TSIR model. Taking the log of both sides of TSIR model (Equation 1), we have

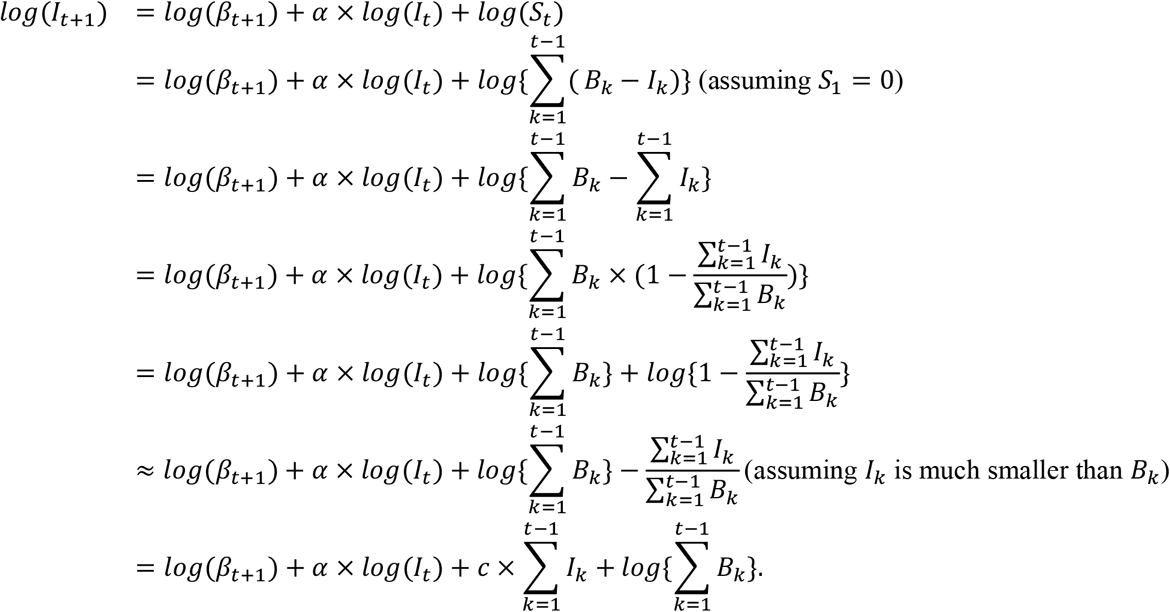

Note that, under the TSIR, we have *α* ≥ 0 (and slightly less than 1) and *c* ≤ 0, which respectively indicate positive association and negative association of the corresponding lagged incidence *I*_*k*_ with the current incidence. The negative association indicated by the parameter *c* may implicitly capture the impact of susceptible depletion. Should the LASSO model (Equation 4) resemble the TSIR, we would expect to see a tendency towards positive coefficients *ψ*_*J*_ associated with the most recent lagged incidence (corresponding to the positive *α* value) and non-positive coefficients at other lagged incidence (corresponding to the non-positive *c* value). Also note that historical births may be absorbed in the intercept term of the LASSO model.

We stress that we are not aiming to draw close equivalence between the TSIR and our LASSO model. Instead, this framing aims to provide some insights into the question that how these two modeling approaches may be interconnected heuristically.

## Data Availability

All data will be made available at https://github.com/msylau/

## 1 Supplemetary Figures 1

**Fig. S1:**
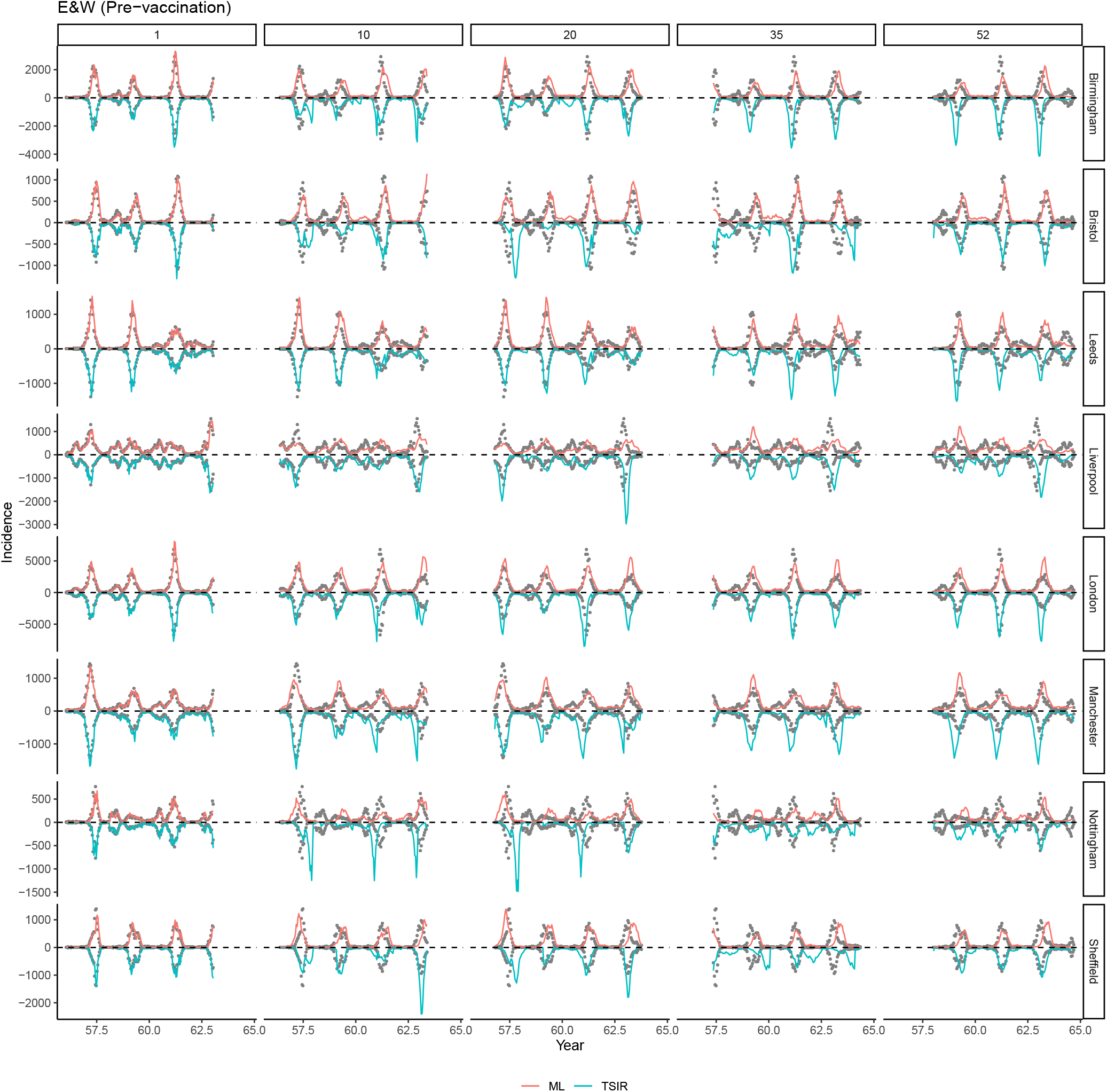
A subset of 1 to 52^*th*^-biweek ahead *out-of-sample* (i.e. period excluding data in the training set) predictions from our LASSO model and the TSIR model, for pre-vaccination measles epidemics in 8 places whose the average population sizes are greater than the critical community size 300,000 from 1944-64.

**Fig. S2:**
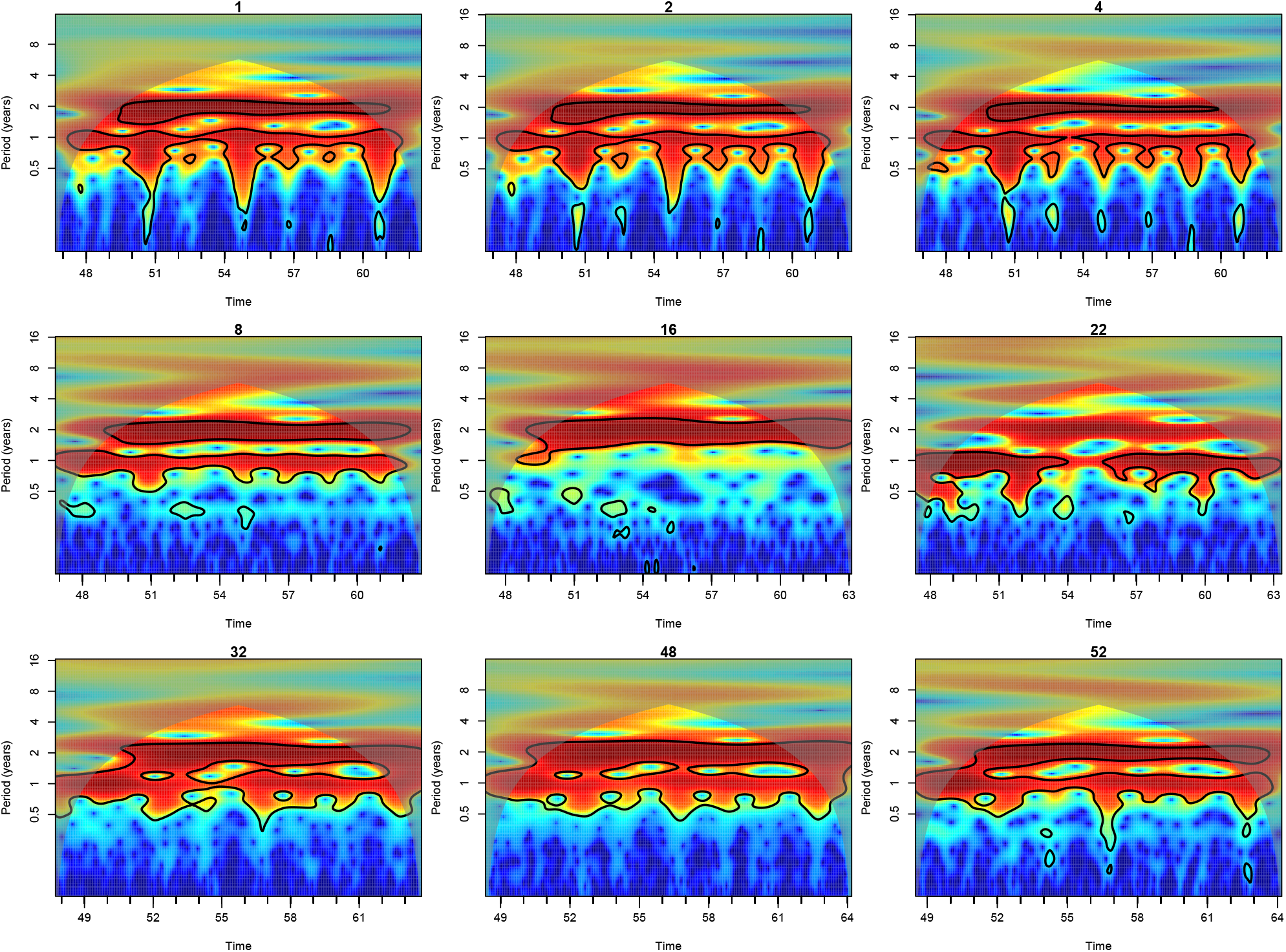
Power spectra of the out of sample predictions for measles in London using the LASSO model (without using birth data in the LASSO model).

**Fig. S3:**
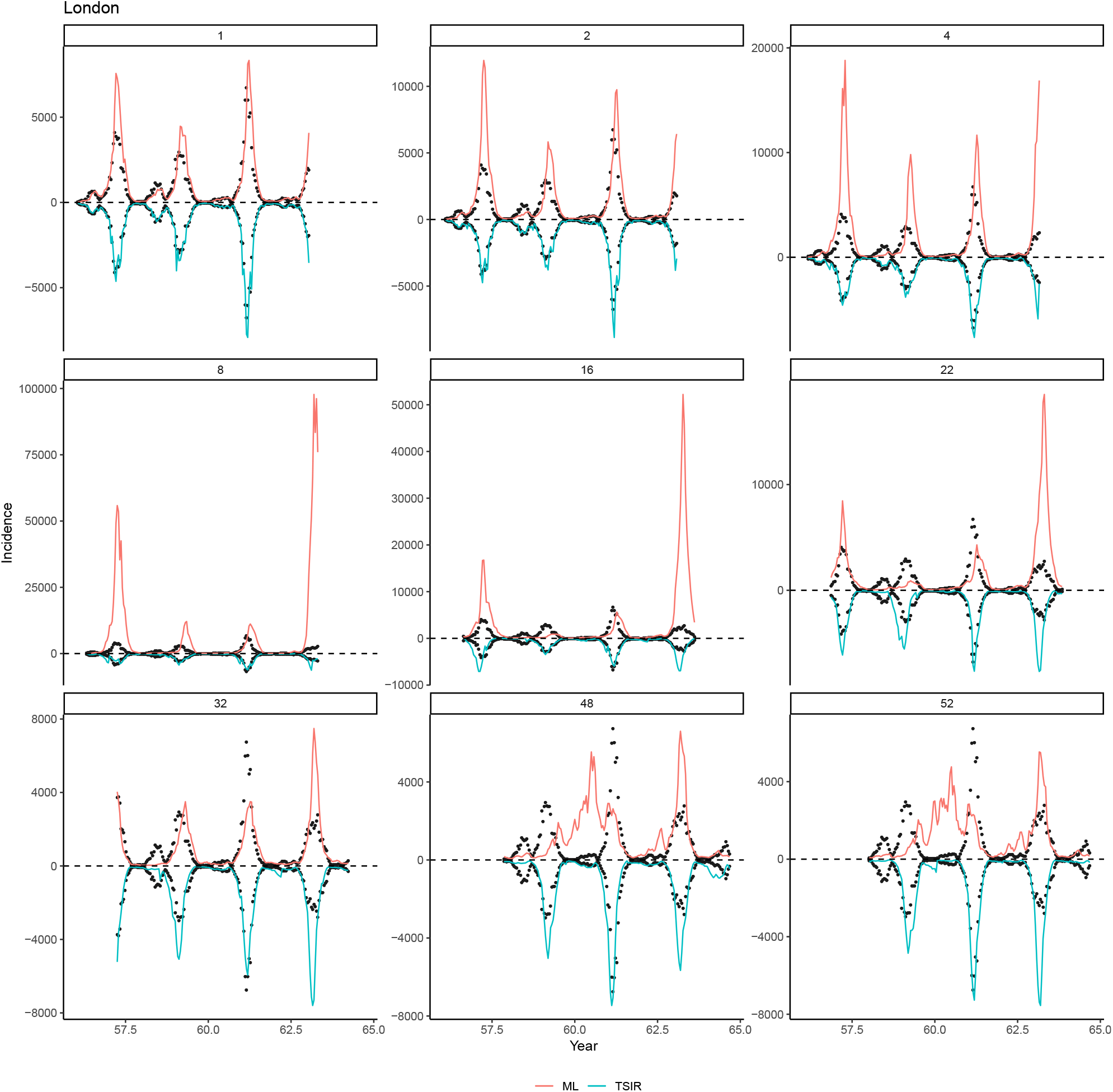
A subset of 1 to 52^*th*^-biweek ahead *out-of-sample* (i.e. period excluding data in the training set) predictions from our LASSO model and the TSIR model, for pre-vaccination measles epidemics in London from 1944-64. Data between 1944-51 (*from London only*) are used to train the LASSO models.

**Fig. S4:**
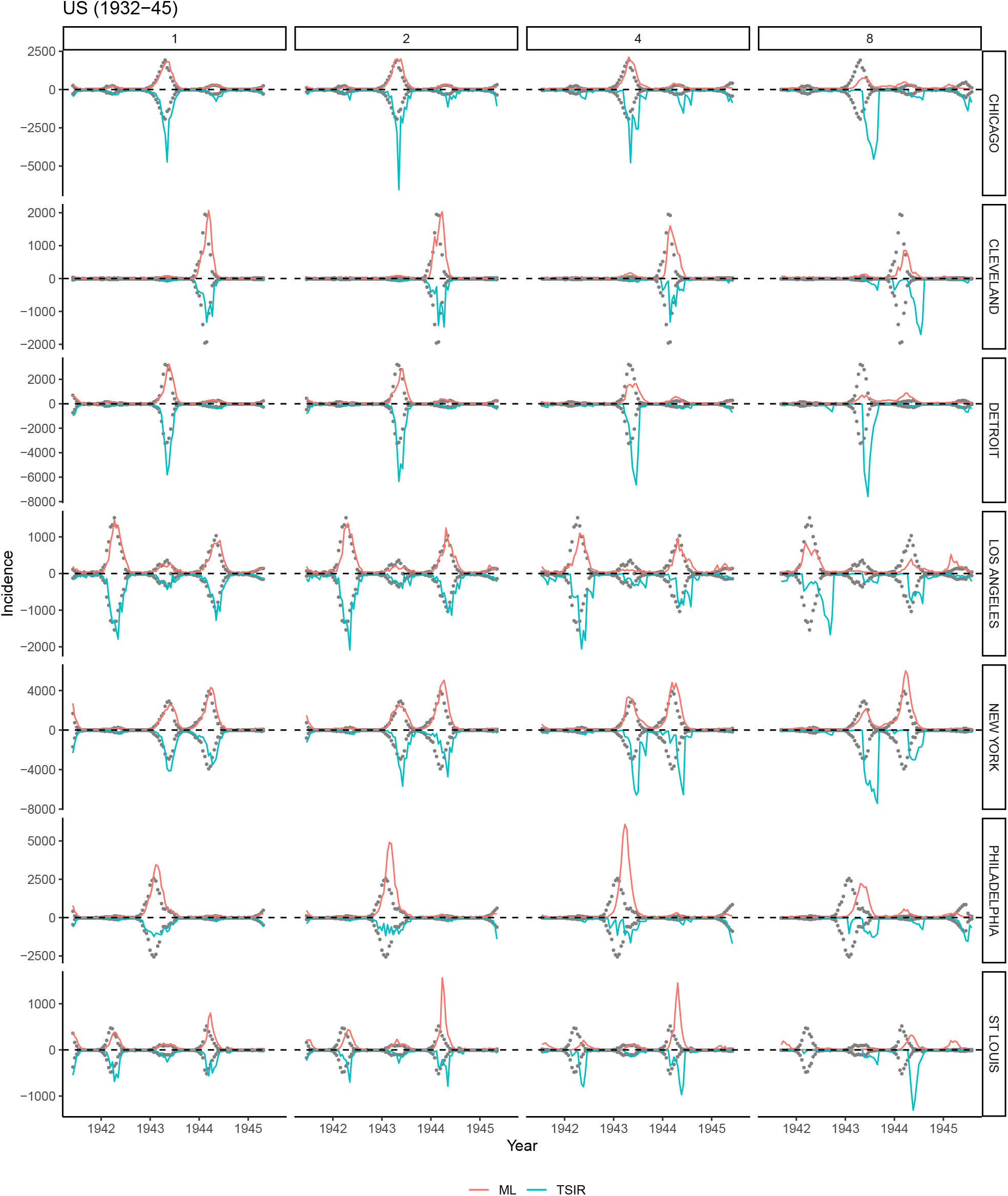
A subset of 1 to 8^*th*^-biweek ahead out-of-sample predictions from our LASSO model and the TSIR model, for measles epidemics in 7 major cities in US from 1932-45. Data between 1932-40 are used to train the LASSO models.

**Fig. S5:**
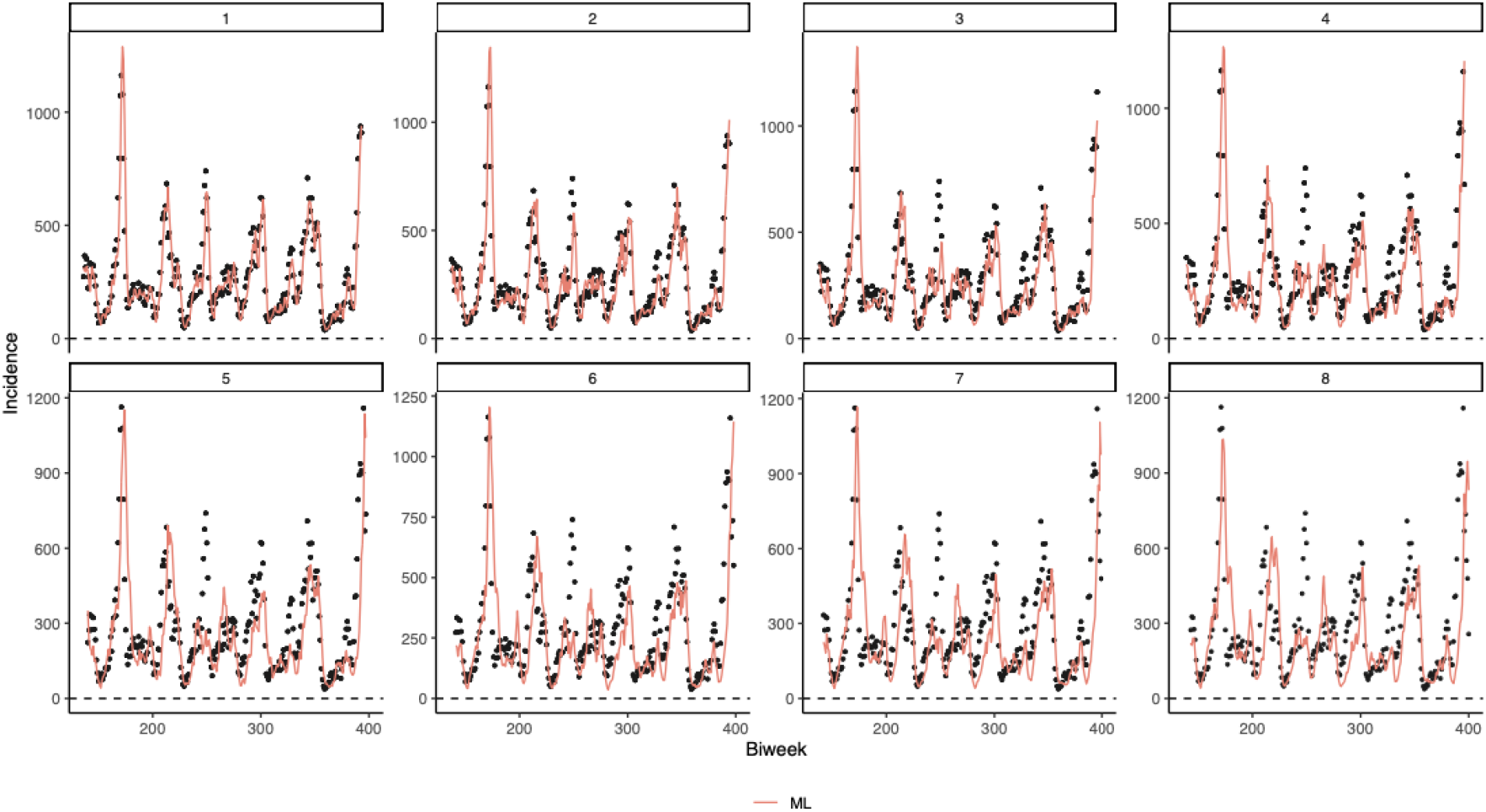
Simulation studies. Our LASSO model is fitted to epidemics generated from a TSIR model. Specifically, local dynamics are simulated from the estimated TSIR model using the pre-vaccination E&W dataset. Using first half of the data for training, our LASSO model can reasonably well predict the outbreak trajectory.

